# A Framework for Analytical Validation of Genomic DNA-Based Shotgun Metagenomic Next-Generation Sequencing Workflows

**DOI:** 10.64898/2026.09.15.26363106

**Authors:** Maurice Chan, Toh Kai Yee, Kimberly Foo, Netra Saravanan, Toh Tzi Shin, Lorraine Lim, Yok-Teng Chionh, Jia-hao Zhang, Wu Mengchu

## Abstract

**Background:** Metagenomic next-generation sequencing (mNGS) is increasingly adopted as a rapid and unbiased alternative to culture-based pathogen detection. However, standardized approaches for analytical validation of mNGS workflows are lacking. Validation strategies based solely on comparison with culture are inherently limited due to low diagnostic yield of culture, and the inability to derive analytical parameters such as limit of detection (LoD) from clinical samples with unknown microbial loads. Systematic analytical approaches are therefore required to define mNGS workflow performance independently of clinical comparators. In addition to diagnostics, mNGS is widely applied to microbiome profiling, where preservation of microbial diversity at low DNA inputs is a key performance criterion.

**Materials and Methods:** The analytical performance of the PaRTI-Seq mNGS workflow (Micronbrane, Taiwan), comprising host depletion, DNA extraction, library preparation, and bioinformatics analysis, was evaluated. Library-level analytical sensitivity was assessed using quantified genomic DNA (gDNA) from *Enterobacter hormaechei*, *Candida albicans*, and *Mycobacterium smegmatis*. Workflow-level sensitivity was evaluated using enumerated suspensions of *E. hormaechei*, *Staphylococcus aureus*, *M. smegmatis*, *Candida albicans* and *Aspergillus brasiliensis*. Microbiome performance at low DNA input was evaluated using stool, saliva and gut mock community samples at 50ng, 5ng, and 0.5ng gDNA input and compared with an alternative low-input library preparation workflow.

**Results:** The library preparation module demonstrated analytical sensitivity down to 0.1 pg of *E. hormaechei* and *C. albicans* gDNA, and 1pg for *M. smegmatis* gDNA. Workflow-level LoDs were determined to be 1 CFU for *E. hormaechei*, 272 CFUs for *S. aureus,* 39 CFUs for *A. brasiliensis*, 36 CFUs for *C. albicans* and 3,702 CFUs for *M. smegmatis*. Differences in extraction efficiencies and extracted gDNA qualities appeared to be important determinants of workflow-level LoDs. For microbiome applications, Micronbrane’s Unison Ultra-low DNA input library preparation kit maintained more consistent bacterial diversity indices down to 5ng DNA input for stool and saliva samples, and the gut mock community than the comparator workflow.

**Conclusion:** This study outlines practical and generalizable analytical validation approaches for mNGS library preparation and complete workflows using defined spike-in models, providing a foundation for informed clinical interpretation and implementation of mNGS in diagnostic and microbiome applications.

## INTRODUCTION

Blood culture (BC) remains the cornerstone for the diagnosis of sepsis, despite well-recognized limitations including prolonged turnaround time and reduced sensitivity for fastidious or slow-growing organisms [1] [2] [3]. Metagenomic next-generation sequencing (mNGS) has been proposed as a complementary or alternative diagnostic approach, offering faster turnaround and broader pathogen detection through unbiased sequencing of nucleic acids in clinical specimens [4] [5] [6]. Unlike culture-based diagnostics, mNGS does not rely on organism viability and is therefore considered more “agnostic” in its detection capability. However, this paradigm shift introduces new challenges in assay validation. Conventional clinical validation relies on comparison against a reference method, typically culture. In the context of mNGS, culture is an imperfect comparator due to its low diagnostic yield and inability to detect many clinically relevant organisms.

Moreover, discrepant results between mNGS and culture cannot be meaningfully interpreted without understanding the analytical performance of both methods. Critically, analytical parameters such as the limit of detection (LoD) cannot be inferred from clinical samples alone, as microbial loads in positive specimens are rarely known. Consequently, analytical validation using defined spike-in models becomes essential for establishing the sensitivity and limitations of mNGS workflows. Several large-scale and laboratory-based evaluations of shotgun metagenomics have highlighted both the importance and the inherent complexity of defining analytical sensitivity for mNGS assays. Multi-center benchmarking studies have demonstrated that assay performance varies substantially across laboratories, workflows, microbial classes, and host backgrounds, and that pathogen detection is driven primarily by relative microbe-to-host abundance rather than absolute microbial concentration [7]. Notably, such studies have proposed that laboratory-developed shotgun metagenomic assays intended for clinical pathogen detection should aim to detect microorganisms at approximately 500 CFU/mL (or copies/mL) within a clinically relevant host context, providing a pragmatic sensitivity benchmark rather than a strict analytical threshold [7]. Complementary laboratory validation studies have further shown that organism-specific LoDs can be established using defined spike-in models, while also demonstrating that host nucleic acid burden and sample quality can markedly reduce assay sensitivity [8].

Importantly, these LoD determinations are only technically meaningful for genomic DNA (gDNA)-based shotgun whole-genome sequencing (WGS) workflows, which target intact microbial cells and therefore permit microbial input to be quantified in terms of colony-forming units (CFU). In contrast, cell-free DNA (cfDNA)-based mNGS assays interrogate fragmented microbial nucleic acids released into biofluids, for which microbial genome equivalents cannot be directly related to organism burden, rendering classical LoD concepts inapplicable. Moreover, the concentration of microorganism-derived cfDNA in clinical specimens is largely unknown and is influenced by multiple biological factors. Consequently, analytical LoDs established using purified cfDNA or synthetic reference materials may be difficult to relate to clinical performance.

A molecular diagnostic workflow generally comprises nucleic acid extraction followed by molecular detection. For mNGS, analytical validation should therefore consider (i) the sensitivity of library preparation at the DNA-level, (ii) extraction efficiency for different microorganisms, and (iii) the combined performance of the entire workflow in terms of colony-forming units (CFU) input.

The analytical requirements of mNGS also depend on its intended application. While infectious disease diagnostics emphasize organism-level detection sensitivity, microbiome profiling aims to preserve microbial community structure and relative taxonomic composition. For microbiome applications, analytical validation should therefore assess the robustness of community profiles across varying DNA input amounts, particularly at low inputs where library preparation may introduce compositional bias or loss of diversity.

The objectives of this study are:

1. Compare the analytical LoDs of the mNGS library preparation in terms of DNA input (in pg) using genomic DNA (gDNA) from *Enterobacter hormaechei*, *Candida albicans*, and *Mycobacterium smegmatis*.
2. Determine the LoD of the complete mNGS workflow using enumerated cell suspensions of *E. hormaechei* (representative Gram-negative bacterium), *Staphylococcus aureus* (representative Gram-positive bacterium), *M. smegmatis* (representative mycobacterium), *Candida albicans* (representative yeast) and *Aspergillus brasiliensis* (representative filamentous fungi).
3. Assess the impact of human gDNA on microbial detection.
4. Compare microbiome diversity preservation at low DNA inputs using different library preparation kits.

Collectively, these studies were designed to establish an analytical validation framework for mNGS workflows, recognizing that the critical performance characteristics differ according to the intended application, whether for infectious disease diagnostics or microbiome profiling.

## MATERIALS AND METHODS

### Host Depletion

Host depletion was performed using the Devin filter (Micronbrane Medical, Taiwan). Whole blood samples were loaded into a syringe attached to the filter and gently passed through into a 15-mL conical tube.

### Sample Processing and DNA Extraction

Blood samples were centrifuged at 400 × g for 15 min at room temperature to obtain plasma. Plasma was spiked with known CFUs of microorganisms. Each sample was subjected high speed centrifugation (14,000g). The resultant bacterial pellet was used for gDNA extraction using the Devin Microbial DNA Enrichment Kit (MEK-01-024; Micronbrane) according to the manufacturer’s instructions. Genomic DNA was eluted in elution buffer (EB) and quantified using the Qubit High-Sensitivity DNA Assay (Thermo Fisher Scientific, USA).

### Library Preparation and Sequencing

Library preparation was performed using the Unison Ultra-low DNA NGS Library Preparation Kit (NLK-01-048; Micronbrane). Two microliters of extracted gDNA were used as input. Following tagmentation, libraries were amplified for 18 PCR cycles, purified using SeraMag beads (Cytiva, USA), and eluted in 11 µL MBG water. Libraries were quantified using Qubit and sequenced on Illumina NovaSeq platform by a commercial service provider.

### Bioinformatics Pipeline and Pathogen Calling

Sequencing reads were trimmed using fastp v0.23.2 2 [9]. Human reads were removed by alignment to the GRCh38 reference genome using bwa-mem (bwa v0.7.17) [10]. Remaining reads were aligned to a curated microbial database comprising approximately 1,400 representative genomes.

Microbial abundance was reported as both percentage of classified microbial reads and reads per million (RPM). A no-template control (NTC) was included in each batch. A microorganism was considered positive if it met both criteria: ≥4-fold enrichment compared with NTC and RPM ≥5.

### Analytical Sensitivity of Library Preparation

Serial dilutions of quantified gDNA were used to determine the analytical sensitivity of the library preparation module. The LoD was defined as the lowest DNA input yielding a positive call.

### Analytical Sensitivity of the Complete Workflow

Enumerated microbial suspensions were serially diluted in saline or host-depleted blood (Supplementary Fig. S1). Plate counts were performed in triplicate. Parallel samples were subjected to DNA extraction, library preparation, and sequencing.

### DNA Input for Microbiome Applications

Stool and saliva samples were extracted using the EZ2 Automated Nucleic Acid Extractor (Qiagen) and normalised to 50 ng, 5 ng, and 0.5 ng total DNA input using RNAse-free water. Each sample was prepared with two different library preparation kits (NEBNext Ultra DNA Library Prep Kit for Illumina and UNISON Ultralow DNA NGS Library Preparation Kit). Zymobiomics Gut Microbiome Standard (D6331) was also extracted and normalised to the same DNA input amounts and run on both library prep kits. The results from each were compared to the expected relative abundance provided by Zymobiomics.

Shotgun metagenomic sequencing was carried out on an Illumina NovaSeq Platform at sequencing depth of 6Gb. Raw reads were quality-filtered using Kneaddata (v0.12.0). Taxonomic profiling was conducted using Metaphlan4 (v4.1.1) Analytical suitability was defined as preservation of expected microbial diversity across DNA input amounts.

All downstream analyses were performed using R (v4.6.0). Alpha diversity was calculated using the Shannon Diversity Index. Beta diversity was assessed using Bray-Curtis dissimilarity computed on relative abundance data and visualised using principal coordinates analysis (PCoA). Taxonomic composition analyses were conducted at species and genus level, with the top 20 most abundant taxa at each level selected for visualisation.

## RESULTS

### Analytical Sensitivity of Library Preparation

The analytical sensitivity of the mNGS library preparation module was first evaluated using serial dilutions of quantified *E. hormaechei* gDNA to define the minimum DNA input that could be detected. The gDNA concentration was determined by fluorometric quantification (Qubit) and diluted across multiple orders of magnitude before being processed through library preparation and sequencing under standard conditions. Detection of *E. hormaechei* was assessed using predefined pathogen-calling criteria applied consistently across all input levels.

Using this approach, the mNGS library preparation successfully detected *E. hormaechei* gDNA inputs as low as 0.1 pg, corresponding to approximately 10 genome copies (Fig. 1). At this input level, *E. hormaechei* reads were consistently identified above background, meeting the established thresholds for positive detection. As expected, decreasing gDNA input was associated with a proportional reduction in organism-specific reads (R² = 0.957), while positive calls were maintained down to the lowest DNA input tested.

**Fig 1.**
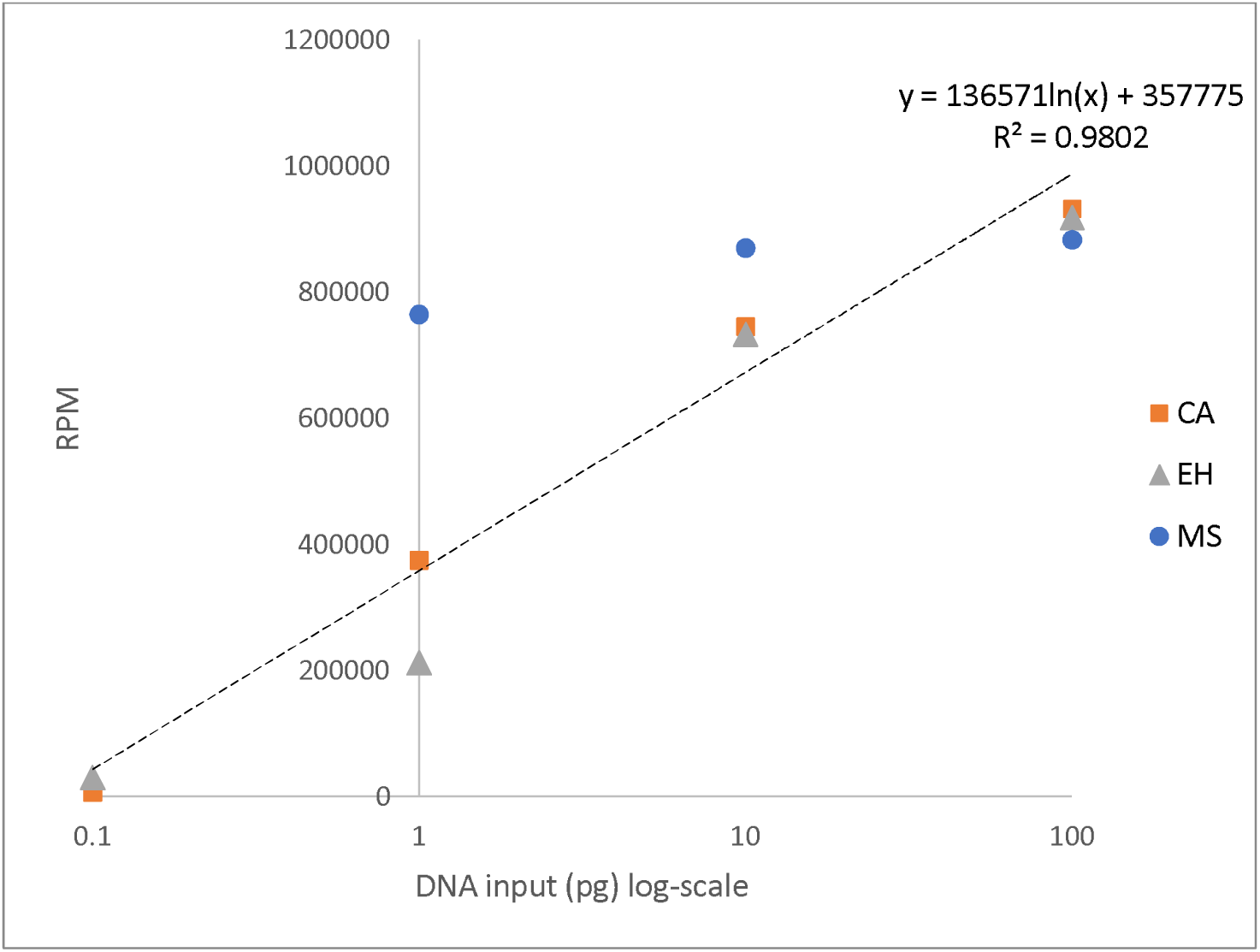
Analytical sensitivity of the mNGS library preparation module using purified gDNA. Relationship between gDNA input and RPM following library prep and sequencing for *E. hormaechei* (EH), *C. albicans* (CA), and *M. smegmatis* (MS) gDNA. All RPM values demonstrated significant fold-change over NTC and were positively called by the analysis pipeline.

To investigate whether library preparation performance was comparable across different microorganisms, serial dilutions of purified gDNA from *C. albicans* and *M. smegmatis* were similarly evaluated. Interestingly, *C. albicans* gDNA demonstrated analytical performance closely resembling that of *E. hormaechei*, with successful detection down to 0.1 pg input DNA. Furthermore, plots of DNA input (pg) against normalized read counts (RPM) exhibited similar linear relationships for both organisms (Fig. 1), suggesting comparable library preparation efficiencies. In contrast, *M. smegmatis* gDNA was only consistently detected down to 1 pg input, with no reads detected at 0.1 pg (Fig. 1). The relationship between input gDNA and resultant RPM was also less proportional for *M. smegmatis*. As extraction was excluded from this experiment, these findings suggest that the purified *M. smegmatis* gDNA itself may have characteristics, potentially including differences in DNA quality or molecular form, that render it less amenable to efficient library construction.

Collectively, these results demonstrate that the library preparation module itself exhibits high analytical sensitivity, approaching single–tens of genome copy detection under controlled conditions. While *E. hormaechei* and *C. albicans* exhibited comparable library preparation performance, the reduced detectability of *M. smegmatis* suggests that genomic DNA from certain microorganisms may be intrinsically less amenable to library construction. Importantly, this analytical assessment isolates library preparation performance from upstream variables such as DNA extraction efficiency and host background, thereby establishing an upper bound for achievable sensitivity within the complete mNGS workflow.

### Analytical Sensitivity of the Complete Workflow

The analytical sensitivity of the complete mNGS workflow was evaluated using enumerated bacterial suspensions to define workflow-level limits of detection (LoD) in terms of colony-forming units (CFU) per sample. Detection was assessed using predefined pathogen-calling criteria, and the LoD was defined as the lowest input CFU at which the organism could be consistently detected.

For *E. hormaechei*, gDNA extracted from the two highest bacterial inputs was quantifiable using Qubit, whereas DNA recovered from lower input samples was below Qubit’s limit of detection (Table 1). In contrast, DNA extracted from *S. aureus* remained below the detection limit of the Qubit assay even at the highest bacterial input tested (Table 2). Despite this, mNGS successfully detected organism-specific sequences from samples in which the extracted gDNA could not be detected by Qubit, demonstrating that library preparation and NGS were capable of detecting microbial gDNA at quantities below the sensitivity of conventional fluorometric DNA quantification.

**Table 1.** Table of NGS results at different CFU input of E. hormaechei cells in (a) saline (E1 to E9) and (b) blood (BE2, BE4, BE7 and BE8). Fold change is defined as RPM_SAMPLE_: RPM_NTC_.

| Sample name | Dilution | CFU in sample (2ml) | Pellet visible? | Qubit after DNA extraction (ng/ul) | Total yield in 80ul of EB (ng) | Total GE | Extraction efficiency | Library prep Qubit (ng/ul) | *E. hormaechei* reads | RPM | Fold change |
| --- | --- | --- | --- | --- | --- | --- | --- | --- | --- | --- | --- |
| **(a) Saline** |  |  |  |  |  |  |  |  |  |  |  |
| E1 | 1.00E-01 | 142,000,000 | Y | 3.68 | 184.00 | 36,269,896 | 25.54% | 3.380 | 8,087,170 | 979,173 | 1181.15 |
| E2 | 1.00E-02 | 14,200,000 | Y | 0.518 | 25.90 | 5,105,382 | 35.95% | 1.980 | 6,476,929 | 972,301 | 1172.86 |
| E3 | 1.00E-03 | 1,420,000 | N | TLTD | NA | NA | NA | 0.398 | 1,385,063 | 826,133 | 996.54 |
| E4 | 1.00E-04 | 142,000 | N | TLTD | NA | NA | NA | 0.804 | 67,312 | 105,682 | 127.48 |
| E5 | 1.00E-05 | 14,200 | N | not done | NA | NA | NA | 0.470 | 32,697 | 29,752 | 35.89 |
| E6 | 1.00E-06 | 1,420 | N | not done | NA | NA | NA | 0.266 | 3,933 | 10,980 | 13.24 |
| E7 | 1.00E-07 | 142 | N | not done | NA | NA | NA | TLTD | 141 | 3,719 | 4.49 |
| E8 | 1.00E-08 | 14 | N | not done | NA | NA | NA | 0.168 | 785 | 26,963 | 32.52 |
| E9 | 1.00E-09 | 1 | N | not done | NA | NA | NA | 0.192 | 670 | 6,039 | 7.28 |
| E-NTC | NTC | 0 | N | TLTD | NA | NA | NA | TLTD | 60 | 829 | 1.00 |
| **(b) Blood** |  |  |  |  |  |  |  |  |  |  |  |
| BE2 | 1.00E+02 | 14,200,000 | Y | TLTD | NA | NA | NA | 4.74 | 1,069,682 | 101,551 | 634.69 |
| BE4 | 1.00E+04 | 142,000 | Y | TLTD | NA | NA | NA | 0.478 | 68,821 | 5,202 | 32.51 |
| BE7 | 1.00E+07 | 142 | Y | TLTD | NA | NA | NA | 0.492 | 334 | 68 | 0.43 |
| BE8 | 1.00E+08 | 14 | Y | TLTD | NA | NA | NA | 0.362 | 927 | 128 | 0.80 |
| BE-NTC | NTC | 0 | Y | TLTD | NA | NA | NA | TLTD | 139 | 160 | 1.00 |

**Table 2.**
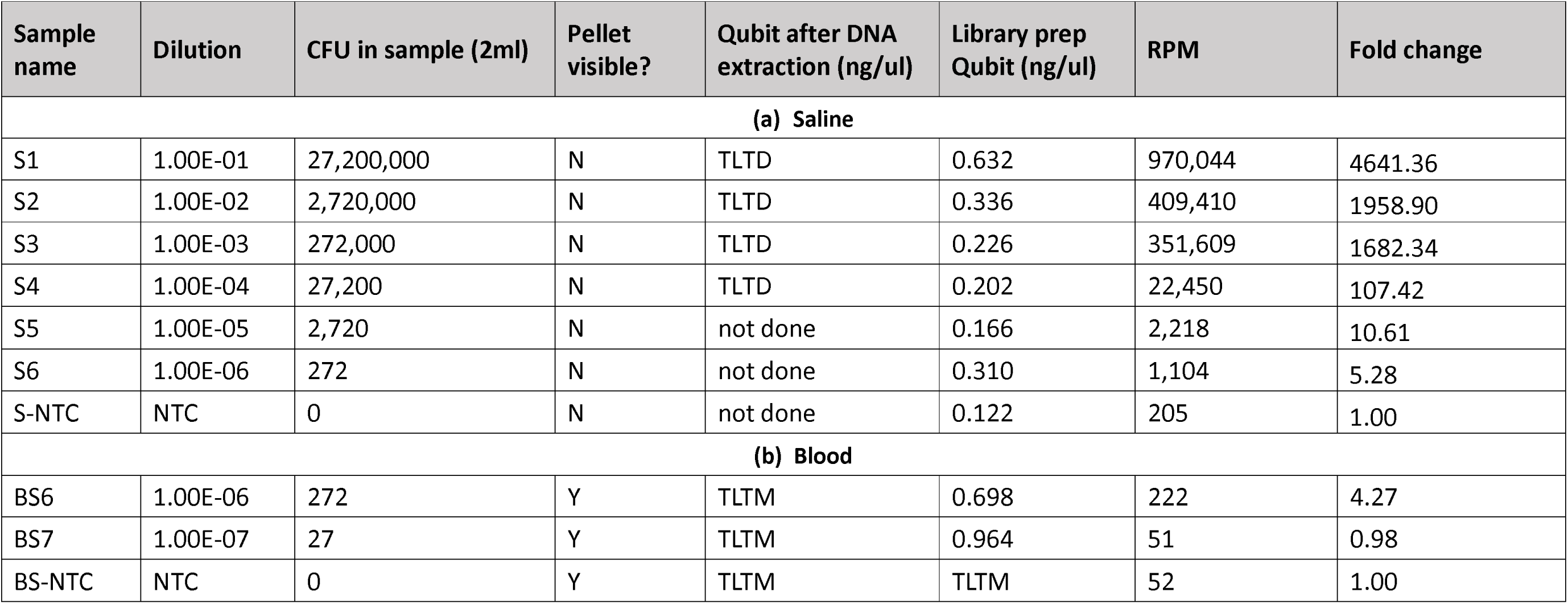
Table of NGS results at different CFU input of S. aureus cells in (a) saline (S1 to S6) and (b) blood (BS6 and BS7). Fold change is defined as RPM_SAMPLE_: RPM_NTC_.

| Sample name | Dilution | CFU in sample (2ml) | Pellet visible? | Qubit after DNA extraction (ng/ul) | Library prep Qubit (ng/ul) | RPM | Fold change |
| --- | --- | --- | --- | --- | --- | --- | --- |
| **(a) Saline** |  |  |  |  |  |  |  |
| S1 | 1.00E-01 | 27,200,000 | N | TLTD | 0.632 | 970,044 | 4641.36 |
| S2 | 1.00E-02 | 2,720,000 | N | TLTD | 0.336 | 409,410 | 1958.90 |
| S3 | 1.00E-03 | 272,000 | N | TLTD | 0.226 | 351,609 | 1682.34 |
| S4 | 1.00E-04 | 27,200 | N | TLTD | 0.202 | 22,450 | 107.42 |
| S5 | 1.00E-05 | 2,720 | N | not done | 0.166 | 2,218 | 10.61 |
| S6 | 1.00E-06 | 272 | N | not done | 0.310 | 1,104 | 5.28 |
| S-NTC | NTC | 0 | N | not done | 0.122 | 205 | 1.00 |
| **(b) Blood** |  |  |  |  |  |  |  |
| BS6 | 1.00E-06 | 272 | Y | TLTM | 0.698 | 222 | 4.27 |
| BS7 | 1.00E-07 | 27 | Y | TLTM | 0.964 | 51 | 0.98 |
| BS-NTC | NTC | 0 | Y | TLTM | TLTM | 52 | 1.00 |

For *E. hormaechei*, the workflow LoD was determined to be 1 CFU per sample (Table 1). This finding indicates that, when extraction efficiency is favorable, the overall workflow can achieve near single-genome copy sensitivity, consistent with the high analytical sensitivity observed at the library preparation level. Although DNA extracted from lower-input samples was not measurable by Qubit, successful pathogen detection by mNGS confirmed that sufficient genomic DNA remained available for library construction and sequencing.

In contrast, the workflow LoD for *S. aureus* was higher, at 272 CFU per sample (Table 2). This may not be attributed to limitations in library preparation sensitivity, but more likely primarily driven by reduced efficiency during DNA extraction. The markedly poorer DNA recovery from *S. aureus*, evidenced by the absence of Qubit-detectable DNA even at the highest bacterial input, further supports this inference. These results highlight pronounced organism-dependent variability in extraction efficiency and demonstrate that workflow-level sensitivity is not uniform across bacterial species.

### Expanded Organism Testing

To assess the applicability of the analytical validation framework across a broader range of microorganisms, the mNGS workflow was further evaluated using fungal and mycobacterial organisms with distinct cellular structures and genome characteristics.

Extraction of the highest fungal and mycobacterial inputs resulted in gDNA concentrations that were readily detectable using Qubit (Table 3a), confirming measurable DNA recovery from all three organisms. Estimated extraction efficiencies ranged from 2.82% to 4.62%, demonstrating that genomic DNA could be recovered despite the distinct cellular structures of the organisms tested.

**Table 3.** Extraction (a) and NGS (b) results of A. brasiliensis, C. albicans, and M. smegmatis spiked into saline or host-depleted blood at known CFUs per sample.

| Sample for Devin extraction | Qubit (ng/ul) | Total ng | Total copies extracted | Input | Efficiency |
| --- | --- | --- | --- | --- | --- |
| *A. brasiliensis* | 0.196 | 15.68 | 392,000 | 13,900,000 | 2.82% |
| *C. albicans* | 1.05 | 52.5 | 1,680,000 | 36,400,000 | 4.62% |
| *M. smegmatis* | 2.04 | 102.0 | 13,260,000 | 370,000,000 | 3.58% |

(b)
| Organism | Input CFU | Matrix | Human reads | Human (%) | Target reads (RPM) | Fold-change |
| --- | --- | --- | --- | --- | --- | --- |
| *A. brasiliensis* | 38,920 | saline | 66,544 | 1.51 | 837,877 | 5,769.82 |
|  | 3,892 | saline | 111,755 | 2.06 | 669,466 | 4,610.10 |
|  | 389 | saline | 219,543 | 5.81 | 445,016 | 3,064.49 |
|  | 39 | saline | 57,339 | 3.65 | 27,779 | 191.29 |
|  | 38,920 | blood | 1,738,651 | 30.68 | 573,808 | 3,951.38 |
|  | 3,892 | blood | 1,678,261 | 55.96 | 158,333 | 1,090.32 |
| *C. albicans* | 36,400 | saline | 20,226 | 17.92 | 6,547 | 390.93 |
|  | 3,640 | saline | 653,803 | 37.94 | 34,691 | 453 |
|  | 364 | saline | 14,499 | 20.6 | 341 | 11.41 |
|  | 36 | saline | 7,659 | 17.14 | 3,558 | 143.88 |
|  | 36,400 | blood | 2,387,311 | 96.69 | 2,302 | 30.08 |
|  | 3,640 | blood | 6,782,182 | 97.6 | 514 | 6.71 |
| *M. smegmatis* | 37,017 | saline | 532,193 | 58.6 | 5,313 | 242.82 |
|  | 3,702 | saline | 296,118 | 45.12 | 727 | 24.31 |
|  | 370 | saline | 1,384 | 42.95 | Not detected | NA |
|  | 3,702 | blood | 14,391 | 99.91 | Not detected | NA |
|  | 370 | blood | 782 | 97.87 | Not detected | NA |

Subsequent mNGS analysis demonstrated that DNA recovered from *A. brasiliensis* and *C. albicans* was suitable for downstream library preparation and sequencing. Serial dilution experiments enabled determination of workflow-level LoDs of 39 CFU and 36 CFU per sample for *A. brasiliensis* and *C. albicans*, respectively (Table 3b). These findings indicated that both organisms could be detected at low microbial inputs despite relatively modest measured extraction efficiencies.

For *M. smegmatis*, organism-specific sequences were successfully detected down to an input of 3,702 CFU in saline. At lower bacterial inputs, *M. smegmatis* was not detected. This reduced workflow-level detectability was consistent with the poorer performance observed when purified *M. smegmatis* gDNA was evaluated directly during library preparation (Fig. 1). Taken together, these findings suggest that the recovered *M. smegmatis* gDNA may be of a quality or molecular form that is less amenable to efficient library construction, with this effect evident at both the library-preparation and complete-workflow levels.

The influence of residual host DNA was further evaluated by comparing organism detection in saline and host-depleted blood matrices. For all five microorganisms, residual host DNA substantially reduced microbial sequencing reads despite prior host depletion. In several instances, organisms readily detected in saline were no longer detected in host-depleted blood at the same microbial input. These findings demonstrate that residual host DNA remains a major determinant of analytical sensitivity in blood specimens and suggest that workflow-level LoDs established under low-background conditions are unlikely to be retained in clinical blood samples, even following host depletion.

### Preservation of genus-level profiles in fecal and saliva samples and within-sample diversity

Having established analytical performance for pathogen detection, we next evaluated the workflows for microbiome profiling, where the principal analytical objective shifts from organism-specific detection sensitivity to preservation of microbial community structure. NEB and UNISON workflows were evaluated using a defined mock microbial community and biological specimens comprising four fecal and four saliva samples at DNA inputs of 50, 5 and 0.5 ng.

At 50 and 5 ng, both NEB and UNISON workflows retained the characteristic profiles of individual samples and the expected distinction between fecal and saliva communities (Fig 2). Fecal samples were dominated by *Phocaeicola*, *Bacteroides*, *Blautia*, *Faecalibacterium* and *Escherichia*, whereas saliva samples were dominated by *Prevotella*, *Streptococcus*, *Veillonella* and *Neisseria*. At 0.5ng, several low-input libraries showed pronounced shifts in dominant genera or reduced taxon representation, particularly among saliva samples under both workflows. Among fecal samples, UNISON retained profiles closer to the higher-input preparations, whereas some NEB libraries diverged more.

**Fig. 2.**
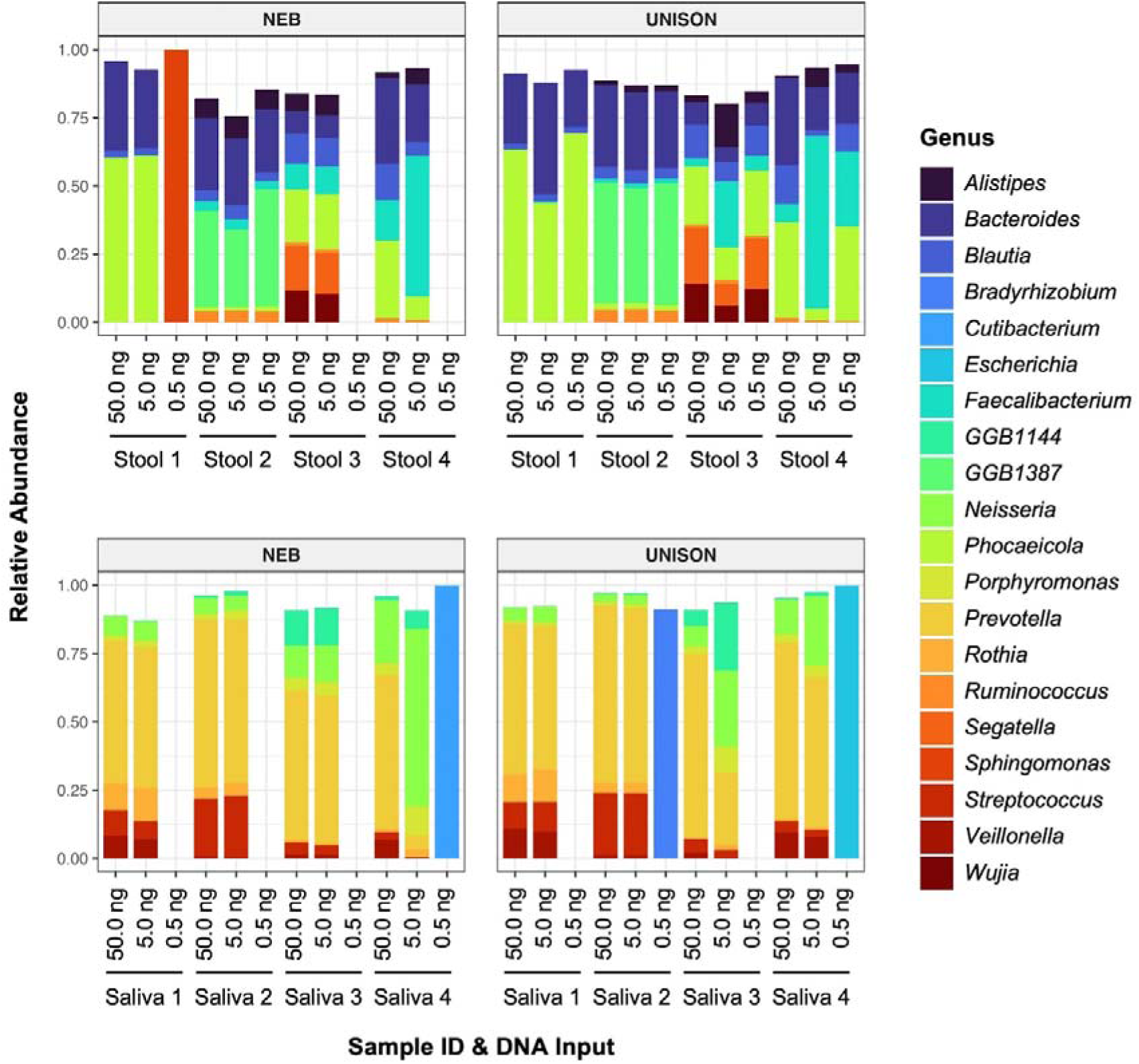
Genus-level compositional consistency across DNA inputs and library-preparation workflows. Relative abundances of the predominant 20 microbial genera in four stool and four saliva samples prepared using the NEB and UNISON workflows at DNA inputs of 50, 5 and 0.5 ng. Each stacked bar represents the genus-level composition of one library, with genera shown according to the legend. Both workflows generally preserved sample-specific profiles at 50 and 5 ng, whereas greater compositional deviations were observed at 0.5 ng in selected samples. Genera not included among the displayed taxa and unclassified reads are not shown; therefore, some bars may sum to less than 100%; missing bars denote unsuccessful library preparation for shotgun metagenomic sequencing.

Shannon diversity was maintained between 50 and 5 ng (Fig 3). UNISON-prepared saliva changed little across these inputs, whereas NEB-prepared saliva showed a modest reduction and greater variability at 5 ng. At 0.5 ng, saliva diversity fell markedly under both workflows, and fecal diversity became lower and more variable, more so for NEB than UNISON.

**Fig. 3.**
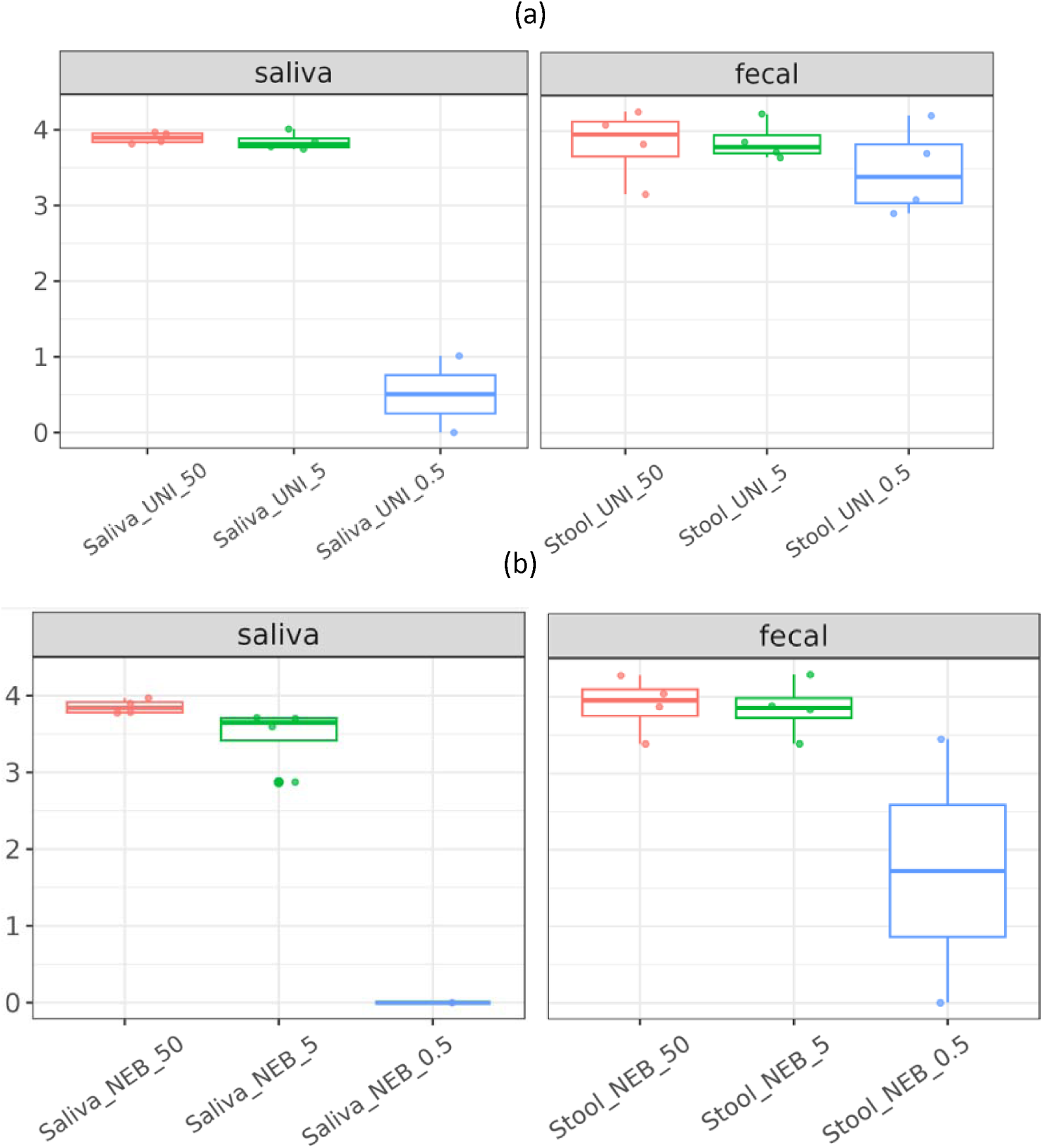
Shannon diversity across DNA inputs and library-preparation workflows. Shannon diversity of fecal and saliva microbial communities prepared using UNISON and NEB workflows at DNA inputs of 50ng, 5ng and 0.5ng. Boxplots showed the median and interquartile range, with individual samples overlaid as points. Shannon diversity was broadly maintained at 50 and 5 ng for both workflows. At 0.5 ng, diversity decreased markedly in saliva samples under both methods and became lower and more variable in fecal samples, with a visually greater reduction under the NEB workflow.

### Preservation of between-sample community structure

Principal coordinate analysis further demonstrated close clustering of 50- and 5-ng libraries within each specimen type, with substantial overlap between NEB- and UNISON-prepared samples (Supplementary Fig S2). This indicates preservation between-sample compositional relationships at these input concentrations. At 0.5 ng libraries showed greater displacement from their corresponding higher-input clusters, most notably among saliva samples; one NEB-prepared fecal sample also showed substantial divergence, whereas the corresponding low-input UNISON fecal profiles were comparatively closer to the main fecal cluster.

### Recovery of the defined mock-community composition

The mock community was used to assess taxon recovery and agreement with manufacturer-specified relative abundances (Supplementary Fig S3). At 50 ng, both workflows produced broadly similar abundance profiles and recovered most taxa present at appreciable theoretical abundance. At 5 ng, the overall community structure remained recognisable, though deviations from the 50-ng profiles became more apparent: UNISON showed limited change for most taxa, whereas NEB profile showed reduced *Veillonella rogosae* and *Fusobacterium nucleatum* and increased *Faecalibacterium prausnitzii* and *Bifidobacterium adolescentis*. Neither workflow reproduced the theoretical composition exactly. Both underestimated *Rosebuira homini* (theoretical 12.43%) and overrepresented *Prevotella corporis* and *Bacteroides fragilis*, and UNISON consistently recovered *B. adolescentis* below expectation. Low-abundance taxa (*Salmonella enterica*, *Methanobrevibacter smithii*, *Enterococcus faecalis*, *Clostridium perfringens*) and the fungal component *Saccharomyces cerevisiae* were undetected or below the visible range. At 0.5 ng, recovery deteriorated and abundances deviated markedly from theoretical values; UNISON retained a broader range of members, whereas the NEB profile was dominated by a limited subset of taxa.

### Overall analytical performance

Taken together, the mock-community and biological-sample analyses showed that NEB and UNISON provided broadly comparable microbiome profiles at 50 ng and generally retained community composition at 5 ng. The defined mock community demonstrated that both workflows were subject to taxon-specific abundance bias, even at higher inputs, and that quantitative agreement with theoretical composition was imperfect. At 0.5 ng, both workflows showed reduced taxon recovery and compositional fidelity, with the greatest deterioration observed in saliva samples and in selected NEB-prepared fecal and mock-community libraries. These findings support 5 ng as a more reliable minimum DNA input for community-level microbiome profiling under the conditions evaluated.

## DISCUSSION

Metagenomic next-generation sequencing (mNGS) encompasses a broad range of clinical and research applications, each with distinct analytical performance requirements. For infectious disease diagnostics, the primary objective is sensitive and accurate pathogen detection, whereas microbiome applications place greater emphasis on preservation of microbial community structure across varying DNA input amounts. Consequently, analytical validation of mNGS workflows should be tailored to the intended application rather than relying on a single validation strategy or performance metric.

An important strength of the analytical framework presented here is the independent evaluation of individual workflow components. Conventional analytical validation studies typically report only the LoD of the complete workflow. Consequently, when an organism is not detected, it is often difficult to determine whether sensitivity is limited by inefficient DNA extraction, poor library preparation performance, or both. By independently characterizing library preparation sensitivity prior to evaluating the complete workflow, the present study demonstrates how the principal analytical bottleneck can be localized to specific workflow components. Such an approach provides greater insight into assay performance while facilitating rational optimization and troubleshooting during assay development. For infectious disease applications, this modular approach enables the respective contributions of library preparation, extraction efficiency, and complete workflow sensitivity to be independently assessed. For microbiome applications, the same framework can be extended to evaluate analytical characteristics more relevant to community profiling, including robustness of taxonomic composition and diversity preservation at low DNA input.

Given the breadth of potential pathogens detectable by shotgun mNGS, it is neither practical nor realistic to determine workflow-level LoDs for every reportable organism. Instead, our findings support the view that comprehensive assessment of organism extractability may provide a more practical analytical strategy than exhaustive LoD determination. In this context, extraction efficiency serves as a useful surrogate for workflow sensitivity, allowing relative detectability across microorganisms to be evaluated without requiring extensive dilution series for every species. However, the present study also demonstrates an important qualification to this approach. While *E. hormaechei* and *Candida albicans* exhibited comparable analytical performance at both the library preparation and workflow levels, *Mycobacterium smegmatis* demonstrated reduced detectability even when purified genomic DNA was subjected directly to library preparation. This observation suggests that genomic DNA quality, or other organism-specific characteristics influencing library compatibility, may not be uniform across microorganisms. Therefore, although comprehensive extractability testing represents a pragmatic and scalable validation strategy, representative assessment of library preparation performance across phylogenetically diverse organisms should also be considered.

An additional conceptual clarification emerging from this work is that classical LoD determination is only applicable to genomic DNA-based shotgun whole-genome sequencing workflows. In these workflows, intact microbial cells or extracted genomic DNA can be quantitatively related to colony-forming units or genome copy input. In contrast, cell-free DNA-based mNGS assays analyse fragmented nucleic acids released into biofluids, for which microbial genome equivalents cannot be directly related to organism burden or extraction efficiency. Applying conventional LoD concepts to cell-free DNA-based mNGS assays may therefore be misleading and highlights the importance of distinguishing between these assay modalities during analytical validation and interpretation.

Rather than viewing analytical validation as determination of a single workflow LoD, we propose that validation should instead be considered as hierarchical characterization of the principal analytical modules that collectively determine assay performance. Based on the findings of this study, technical validation of genomic DNA-based mNGS workflows should comprise three complementary components: (i) determination of library preparation sensitivity using purified genomic DNA, (ii) comprehensive assessment of extraction efficiency across representative microorganisms, and (iii) limited workflow-level LoD determination using enumerated spike-in models in biologically relevant matrices. Such an approach complements clinical validation by providing a more informative understanding of workflow limitations and enabling more meaningful interpretation of discordant results between mNGS and conventional microbiological methods.

While workflow sensitivity is the principal analytical consideration for infectious disease diagnostics, microbiome sequencing places greater emphasis on preserving community composition across varying DNA input amounts. The present study demonstrates that analytical validation for microbiome applications should therefore focus less on organism-specific LoD and more on robustness of diversity indices, taxonomic composition and reproducibility at low DNA input. Together, these findings illustrate that analytical validation strategies should be tailored according to the intended clinical application of mNGS, despite sharing common workflow components.

In conclusion, the analytical validation strategies presented here provide a practical framework for evaluating genomic DNA-based shotgun mNGS workflows for diagnostic and microbiome applications. By independently assessing library preparation, DNA extraction, and complete workflow performance, this modular approach identifies the principal determinants of analytical sensitivity before clinical validation. Such a strategy facilitates workflow optimization, establishes a more informative analytical foundation for clinical implementation, and may contribute to more standardized validation of future mNGS assays.

## Data Availability

All data produced in the present study are available upon reasonable request to the authors.

---

**Supplementary Fig. S1.**
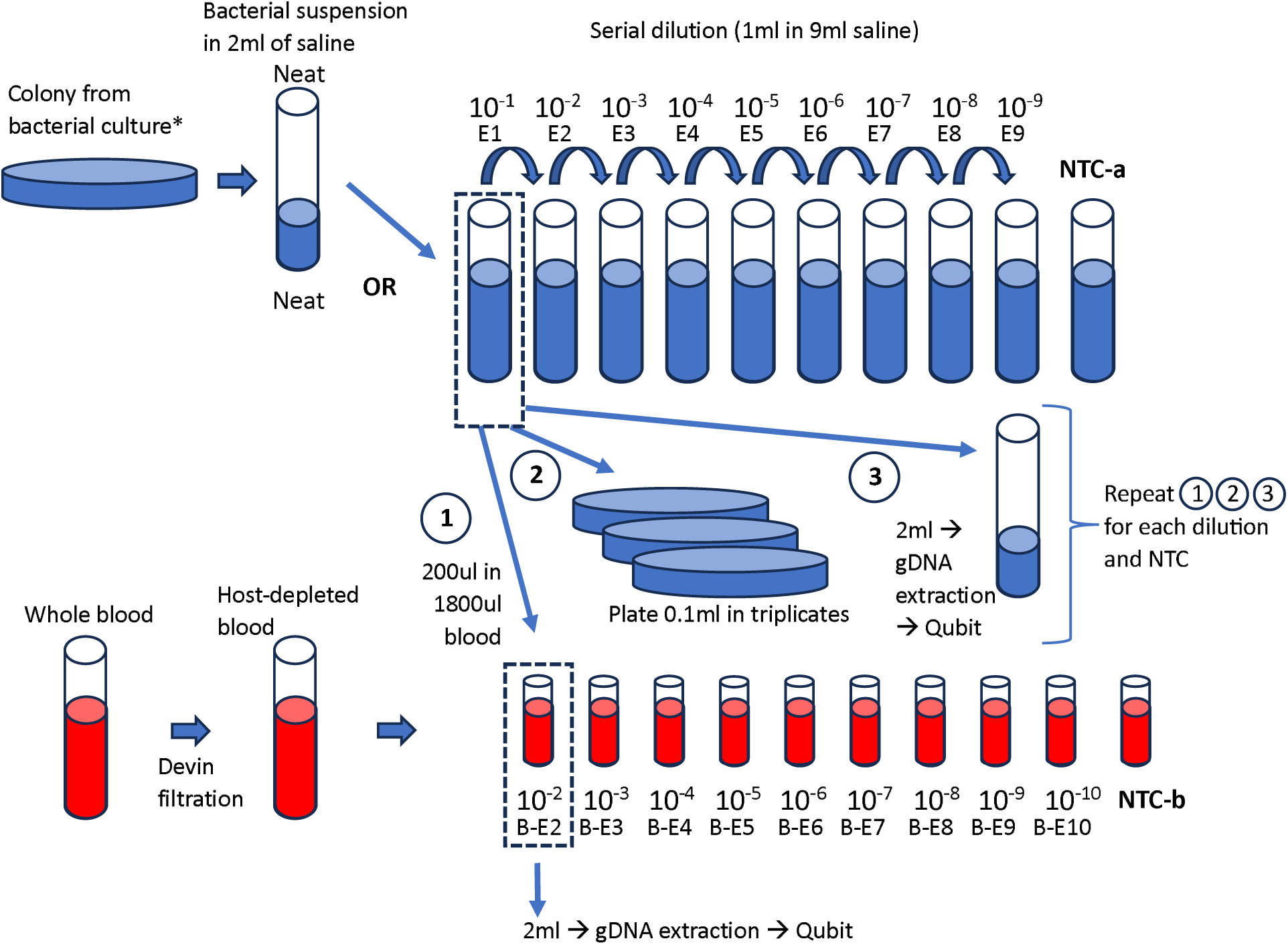
Experimental plan for evaluating the analytical sensitivity of the mNGS workflow using enumerated bacterial suspensions.

**Supplemental Fig. S2.**
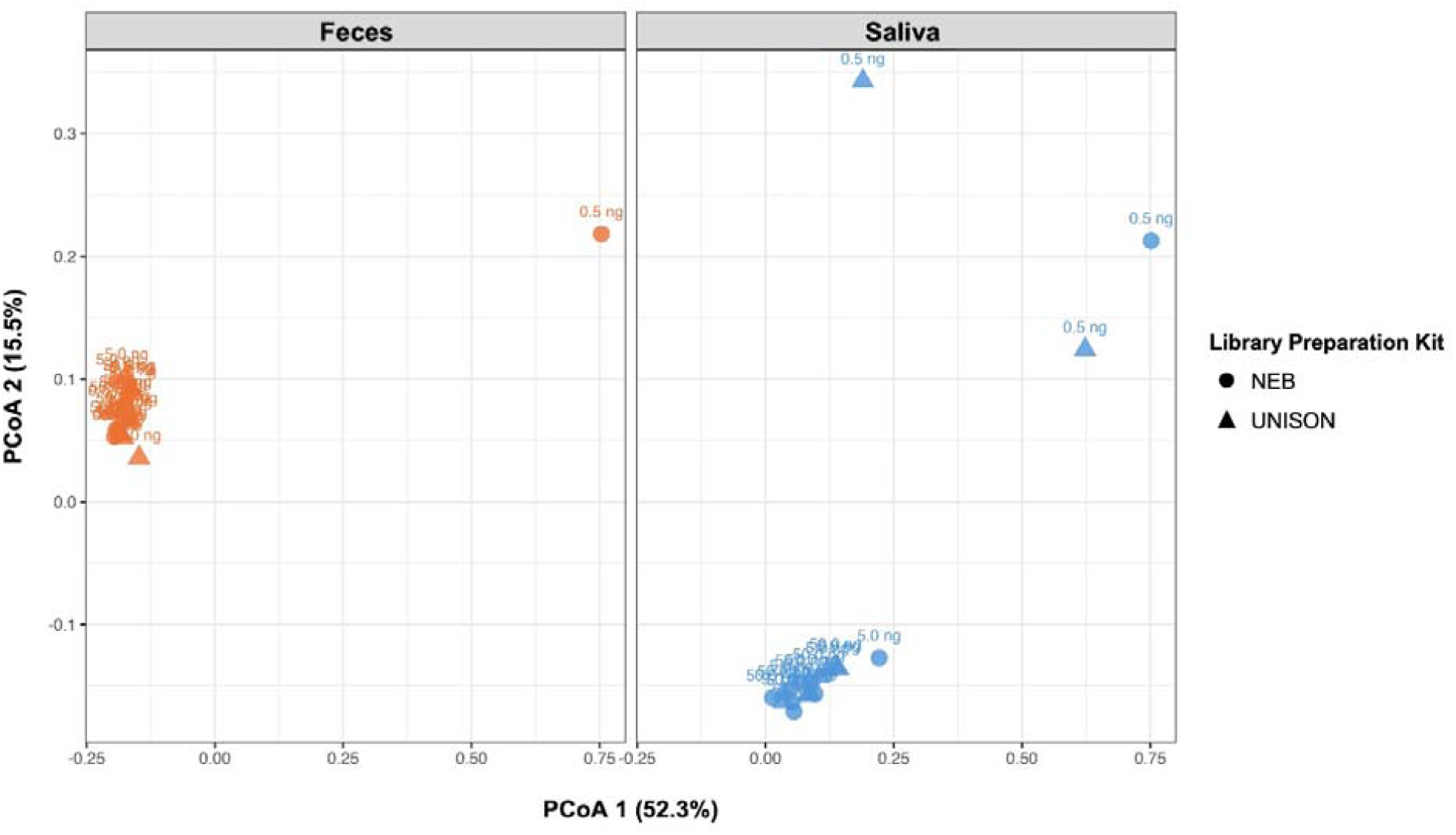
Principal coordinate analysis of microbial community composition based on Bray–Curtis dissimilarity across DNA inputs and library-preparation workflows. PCoA of fecal and saliva microbial profiles prepared using the NEB and UNISON workflows at DNA inputs of 50, 5 and 0.5 ng. Points represent individual libraries, with circles indicating NEB and triangles indicating UNISON. PCoA1 and PCoA2 explained 52.3% and 15.5% of the total variation, respectively. Libraries prepared from 50 and 5 ng DNA clustered closely within each sample type, whereas 0.5 ng libraries showed greater displacement, particularly among saliva samples and one NEB-prepared fecal sample.

**Supplemental Fig. S3.**
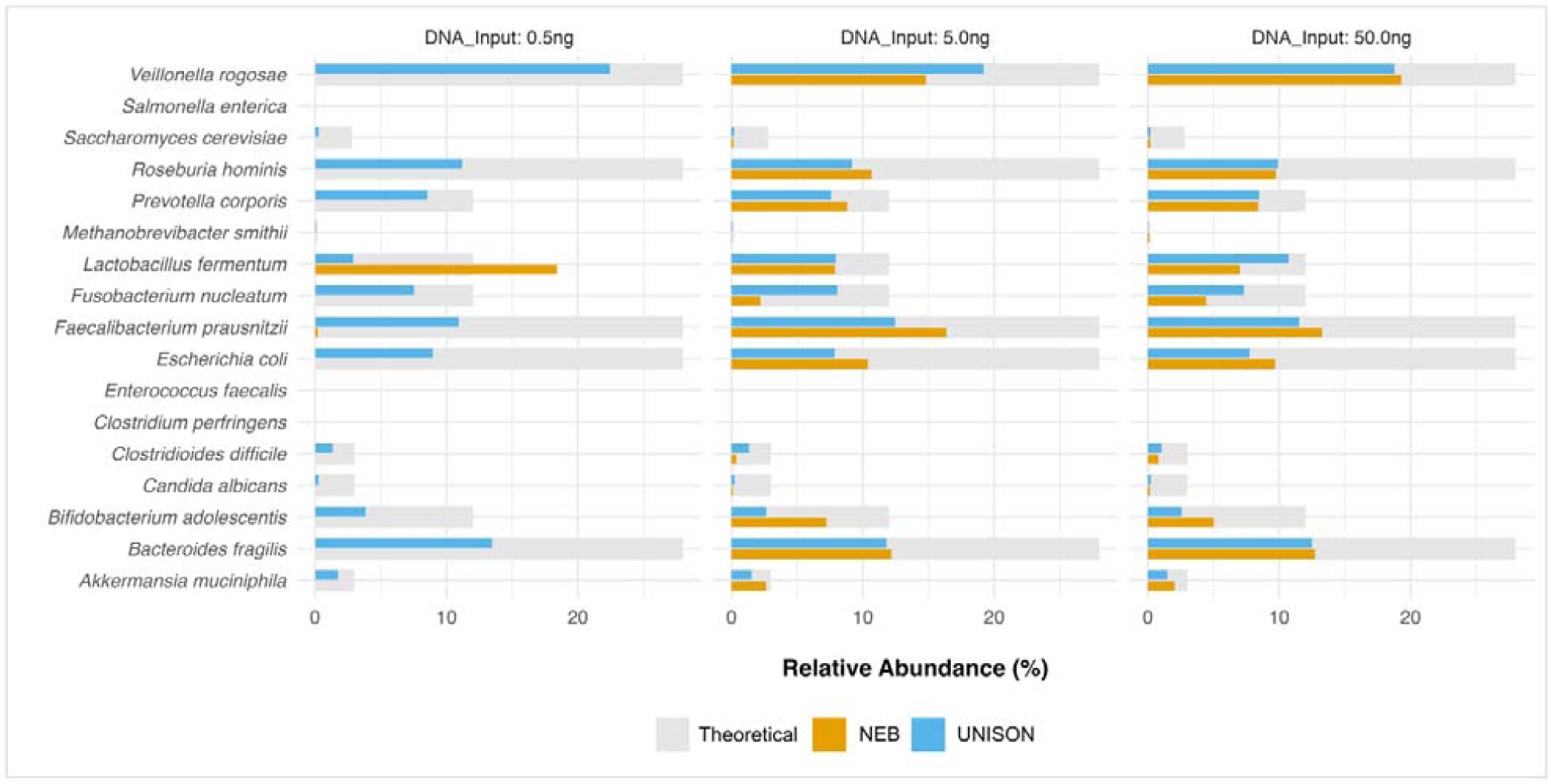
Relative abundance comparison of the Gut Microbiome Standard (D6331) between the NEB and UNISON library preparation kits In the 0.5ng sample, the relative abundance percentage for UNISON follows a similar pattern to that of 5ng and 50ng, except for a decrease in *Lactobacillus fermentum* and an increase in *Veillonella rogosae*. NEB did not successfully capture the Gut Microbiome Standard in the 0.5ng sample as seen in Figure S3, with only *Lactobacillus fermentum* being captured. Beta-diversity was assessed using the Bray-Curtis method; UNISON samples cluster together indicating a high level of similarity, while NEB samples are spread apart in comparison in the PCoA plot.

**Supplemental Figure S4.**
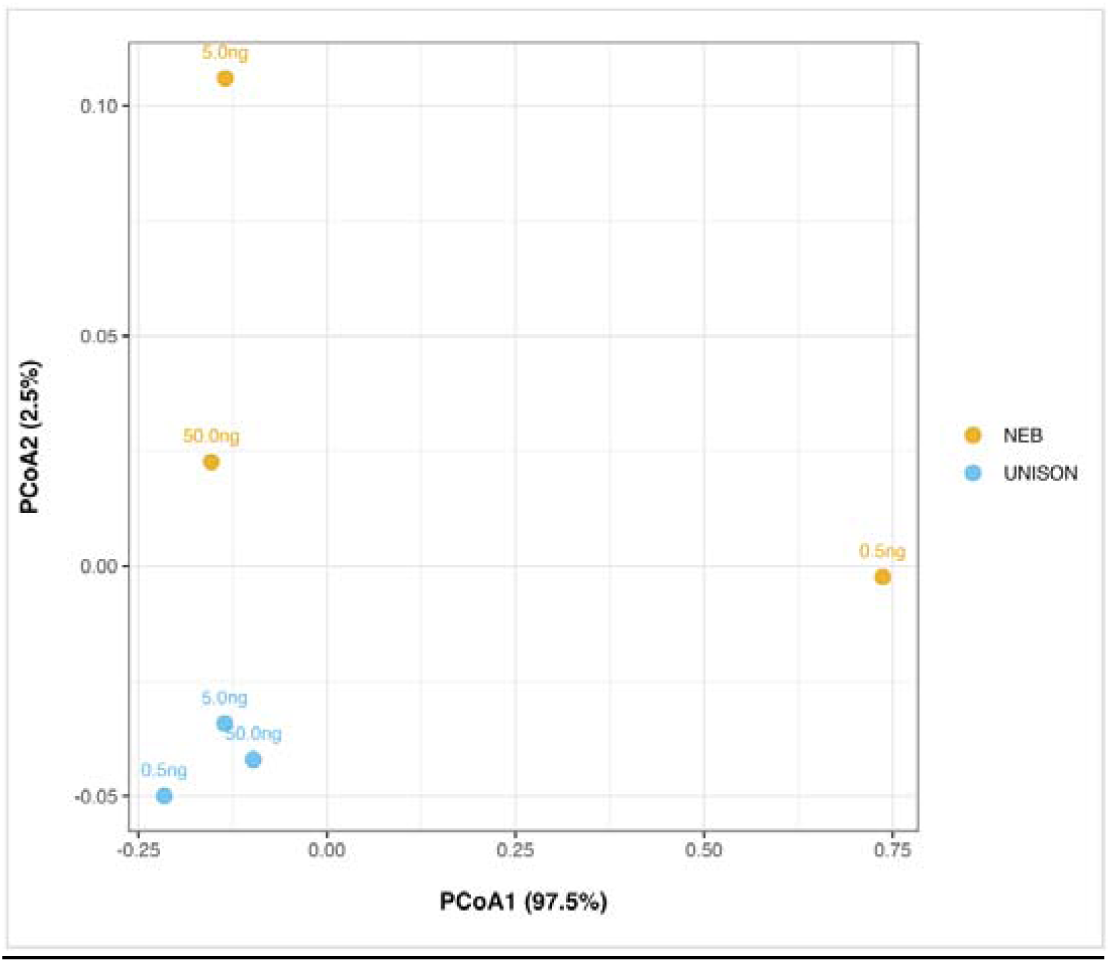
PCoA plot depicting the dis/similarities between sequenced Gut Mock Community with the two library preparation kits at 50ng, 5ng and 0.5ng total input.

